# Adolescent weight control behaviours and adult depressive symptom and body mass index trajectories

**DOI:** 10.64898/2026.07.08.26357532

**Authors:** Bianca Siminea, Ilaria Costantini, Ariana Kular, Gemma Lewis, Glyn Lewis, Francesca Solmi, Madelaine Davies Kellock

## Abstract

**Importance:** In adolescence, attempts to lose weight are common, but their long-term impacts on mental and physical health are not known.

**Objective:** To investigate the association between adolescent dieting and exercising to lose weight and adult trajectories of depressive symptoms and body mass index (BMI).

**Design:** A longitudinal cohort study of children born between April 5 and 11, 1970, and followed up to age 51 years.

**Setting:** Adolescents in the 1970 British Cohort Study in England, Wales and Scotland.

**Participants:** A total of 4,650 adolescents with available exposure data.

**Exposures:** Self-reported lifetime dieting or exercising for weight loss measured at age 16 years.

**Main Outcomes and Measures:** Depressive symptoms measured with the nine-item Malaise Inventory, and BMI derived from self-reported height and weight, at ages 26, 30, 34, 42, 46, and 51 years.

**Results:** Among 4,650 adolescents (56.7% girls, 97.7% White), 1,938 (41.7%) had dieted and 343 (7.4%) had exercised for weight loss by age 16 years. In fully adjusted analyses controlling for a wide range of child- and family-based confounders including prior BMI and emotional difficulties, there was evidence that adolescents who had dieted had higher adult depressive symptom trajectories (adjusted mean difference [aMD] 0.13, 95% CI 0.03-0.24, p=0.015) and higher and increasing adult BMI trajectories than those who had not dieted. There was also evidence that adolescents who exercised for weight loss had higher adult depressive symptom (aMD 0.18, 95% CI 0.02-0.34, p=0.031), and BMI trajectories (aMD 0.37, 95% CI -0.03, 0.78, p=0.071), though evidence of the latter was weak.

**Conclusions and Relevance:** Behaviours aimed at weight loss occurring in adolescence might be a shared risk factor for depressive symptoms and high BMI in adulthood. If causal, these findings could suggest that reducing pressures to lose weight in adolescence may help prevent poor mental and physical health across the lifecourse.

**Key Points:** *Question:* Are dieting and exercising to lose weight in mid-adolescence associated with higher adult trajectories of depressive symptoms and body mass index (BMI)?

*Findings:* Using data from 4,650 adolescents in the 1970 British Cohort Study, adolescents who dieted to lose weight had higher depressive symptoms and higher BMI trajectories from ages 26 to 51. Adolescents who exercised to lose weight had higher depressive symptom, but not BMI, trajectories. These associations were not explained by prior differences in BMI or emotional difficulties in childhood.

*Meaning:* Attempts to lose weight in adolescence may have long-lasting, negative consequences for mental and physical health.

## Introduction

The rising prevalence of depression and overweight or obesity are major global public health challenges, affecting an estimated 332 million^1^ (4.1% of the global population) and 2.1 billion^2^ (26.9%) people globally in 2021, respectively. Depression and overweight or obesity often co-occur across the lifecourse^3^ and, as young adulthood is a key time for the onset of depression^4^ and weight gain^5^, this could suggest they share a risk factor in adolescence.

Attempts to lose weight – such as through dieting and exercising – are common in adolescence, particularly in girls.^6^ Termed weight control behaviours, they may occur in response to body dissatisfaction, exposure to gendered cultural ideals for body weight and shape (broadly, thinness in women and muscularity in men) in media, or after being told to lose weight by a doctor or family member, among other reasons.

As weight control behaviours appear to have been rising over time, independently of increases in BMI^6^, they have been proposed as one of several potential drivers of the increases in depression and BMI seen in the past decades, particularly among girls.

Several longitudinal studies suggest an association between early adolescent weight control behaviours and late adolescent or early adulthood depressive symptoms^7–9^, particularly among girls^7^, as well as BMI^9–13^, though others find no evidence for the latter^14^. However, many of these studies are limited by omitting key confounders, such as prior depressive symptoms^10–12,14^ or parental depressive symptoms and BMI^7–14^. Evidence from a recent genetically informed study suggests a causal association between the cognitive aspects of body dissatisfaction in late adolescence and subsequent depressive symptoms and BMI, albeit to a lesser extent for the latter.^15^ However, this study may have been underpowered to detect small effect sizes and had a relatively short follow up, so it is unclear how these associations may persist over time.

No studies have looked at the long-term impact of weight control behaviours into mid-adult life, which might highlight long-lasting associations through to an age when chronic diseases often emerge, potentially strengthening arguments for prevention. Given the current policy focus on preventing obesity, which often promotes individual diet and exercise changes, it is important to understand the potential for unintended negative consequences on mental health. Therefore, we aimed to investigate the associations of adolescent dieting and exercising to lose weight with trajectories of depressive symptoms and BMI up to age 51 years. Given gendered body ideals and mixed evidence of stronger associations in girls, we also investigated whether associations differed by sex.

## Methods

### Sample

We used data from the 1970 British Cohort Study (BSC70), an ongoing birth cohort study which aimed to recruit all 17,287 children born between April 5th and 11th 1970 across England, Scotland, Wales, and Northern Ireland. Since then, only children born in England, Scotland and Wales, and their parents, have been followed up via questionnaires, medical examinations, and record linkage at ages 5, 10, 16, 26, 30, 34, 38, 42, 46, and 51 years. Sweeps prior to 2000 received ethical approval through an internal review, as was standard practice at the time, and since 2000, from the National Health Service Research Ethics Committee.^16^ Full details of the study can be found elsewhere.^17,18^ We included participants with valid exposure data at age 16. In cases of twins or triplets, we retained one cohort member at random to avoid biases arising from shared genetic and environmental factors.

### Outcomes

#### Depressive symptoms

Every four years from age 26 to 51 (excluding age 38), participants completed the Malaise Inventory, a well-validated scale capturing symptoms of psychological distress and depression.^19^ Whilst at ages 26 and 30 years, the study included the full 24-item scale^20^, after age 34 years, only nine items capturing feelings of depression, fatigue, anxiety, and irritability were retained, which we then used across all time points for consistency (**eTable 1** in the Supplement). Each item was assessed with a binary (no/yes) answer, which we summed to derive a total score ranging 0 to 9, where higher scores indicate greater depressive symptoms.

#### BMI

Participants self-reported their height, in feet and inches or metres and centimetres, and weight, in stones and pounds or kilograms, every four years from age 26 to 51 (excluding age 38). We converted any responses in imperial units into metric and derived BMI using the formula *weight/(height^2^*). At age 46 only, height and weight were also measured by a nurse using a Leicester stadiometer and Tanita BF 522W scales.^21^ We use BMI derived from objectively measured height and weight at age 46 in sensitivity analyses.

### Exposure

#### Weight control behaviours

When participants were around 16 years old, they were asked “Have you ever tried to lose weight or avoid putting on weight?”. Response options were Yes, or No. A follow-up free-text question asked: “If yes, please describe what you did”, from which the BCS70 study team derived two variables indicating whether this was through dieting (yes or no) or exercise (yes or no). From these, we derived two binary variables indicating lifetime dieting or exercising for weight loss if participants had reported lifetime attempts at weight loss and that these had occurred by either of these two methods.

### Confounders

We adjusted for a range of confounders informed by literature- or theory-based assumptions of their association with both exposure and outcome. These were: child’s sex at birth (male or female) and ethnicity (White or minoritised), highest parent education (compulsory or non-compulsory), father’s (or maternal, where missing) occupational class (manual or non-manual), mother’s marital status (married or unmarried), child’s birthweight in kilograms, mother’s age at child’s birth (in five-year bands), mother’s cigarette (never smoked, stopped before pregnancy, smoked in pregnancy) and alcohol (any or none) use in pregnancy, mother’s BMI and depressive symptoms when the child was aged 10, child’s self-esteem at age 10, child’s health-related behaviours at age 10 (smoking (any or none), breakfast consumption, physical activity, television watching (all as never, sometimes, often), and child’s medical conditions (any or none)), objectively measured BMI and emotional symptoms (Rutter Behaviour Scale^22^) at age 10. We controlled for BMI and emotional symptoms at age 10 over age 16 years (i.e. study baseline) to minimise the potential for these to be on the causal pathway between exposure and outcome given the concomitant measurement with the exposure. Although the exposure captures lifetime weight-loss behaviours, which could have started prior to age 16, the prevalence of these is low before early adolescence^23^, hence we assumed that BMI and emotional symptoms at age 10 years would have preceded the exposure for the majority of the sample. See **eMethod1** for full details of confounder measurement.

### Statistical analyses

All analyses were conducted in Stata MP 18.5.^24^ Among participants with valid exposure data, we described the sample characteristics and the distribution of dieting and exercise across confounders using frequencies and proportions. We also compared the characteristics of the sample with valid exposure data to those of the full cohort. We then described frequencies and proportions of missing confounder data and explored differences in characteristics of those with valid and missing outcome data. We imputed missing confounder and outcome data using multiple imputation by chained equations, imputing fifty datasets using all variables included in the main analyses and additional auxiliary variables to improve precision (**eMethod2**).

We used univariable and multivariable multilevel linear regression models with a random intercept on participant and a random slope on linear time to investigate the association between dieting and exercise for weight loss at age 16 and depressive symptom and BMI trajectories from 26 to 51. With age centred at 26 years, we first ran unconditional models using linear and quadratic age indicators to investigate the shape of outcome trajectories over time. We retained both if there was evidence of an association with the outcome and evidence of improved model fit (i.e., lower Akaike Information Criterion).

Subsequently, for each exposure-outcome combination, we ran an unadjusted model followed by multivariable models sequentially adjusting for confounders, as shown in **Table 1**. In fully adjusted models, we investigated whether associations varied by sex and/or over time. First, we included an interaction between sex and exposure and retained it in the model if there was evidence of sex differences, based on p values for the interaction (p ≤0.05). Second, we included an interaction between with linear age and exposure, again retaining based on interaction p value. Finally, where a quadratic age indicator had been retained from unconditional modelling, we included both linear age—exposure and quadratic age—exposure interactions, retaining if their p values were ≤0.05). We plotted the predicted margins and 95% confidence intervals (CI) from fully adjustment models including any interactions retained.

**Table 1.** Model specification for multilevel models.

| **Models** | **Confounders** |
| --- | --- |
| Model 0 | Unadjusted |
| Model 1 | Model 0 + child sex and ethnicity |
| Model 2 | Model 1 + parents' education, father's occupational class, parents' marital status at child's birth |
| Model 3 | Model 2 + mother's age at child's birth, mother's cigarette and alcohol use in pregnancy, child birthweight |
| Model 4 | Model 3 + mother's BMI and depressive symptoms when child aged 10 |
| Model 5 | Model 4 + child's self-esteem, breakfast consumption, physical activity, television habits, smoking, and general health at age 10 |
| Model 6 | Model 5 + child's BMI at age 10 |
| Model 7 | Model 6 + child's emotional symptoms at age 10 |

We ran all multilevel models on a sample of participants with valid exposure and imputed outcome and confounder data, as well as a sample with complete data on exposure and confounders and at least one outcome measure (as multilevel modelling can account for missing outcome data using maximum likelihood estimation).

Given the potential for reporting bias when relying on self-reported BMI, as a sensitivity analysis, we ran univariable and fully adjusted analyses using linear regression (using all confounders in the main analysis) testing the association between each exposure and (i) self-reported BMI at age 46 and (ii) nurse-measured BMI at age 46, comparing estimates to assess whether using measured BMI changed the association. We performed these on a sample with valid exposure data and imputed outcomes and confounders.

## Results

### Sample

Of the 16,672 cohort members who took part in the first sweep of BCS70, 4,650 (27.9% of the total sample and 40% of those invited to the age 16 survey) had data on both exposures. Among participants with exposure data, 2,637 (56.7%) were female, 4,108 (97.7%) were White, and 3,091 (66.9%) had a parent who had attended compulsory education.

Within the analytical sample, 1,938 (41.7%) participants reported having dieted and 343 (7.4%) having exercised to lose weight at some point in their life at age 16 years. Both dieting and exercise were more common among girls and children with an overweight or obese BMI at age 10. Dieting was also more prevalent among children who reported never being physically active or whose mother had overweight or obese BMI (full results in **Table 2**).

**Table 2.**
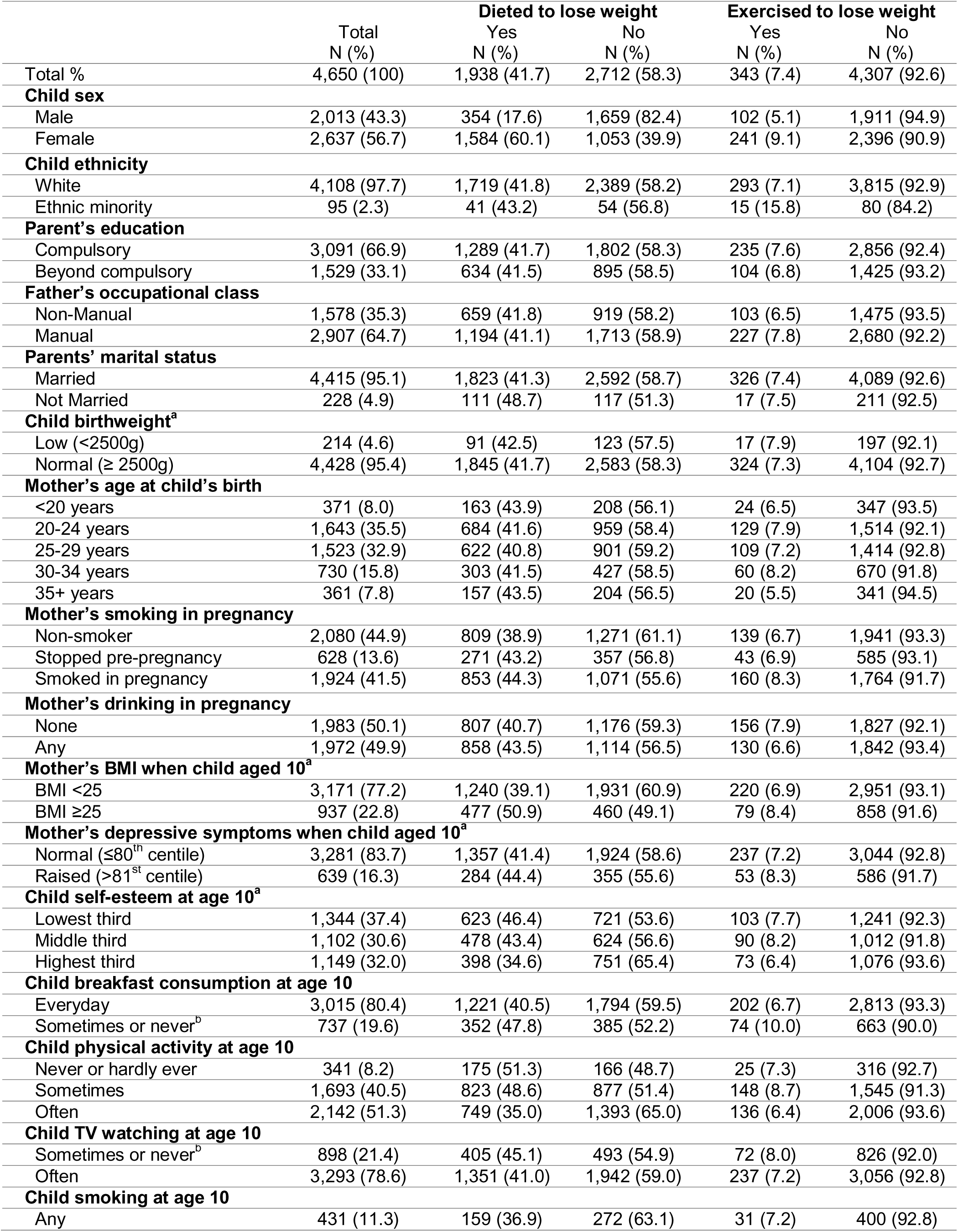
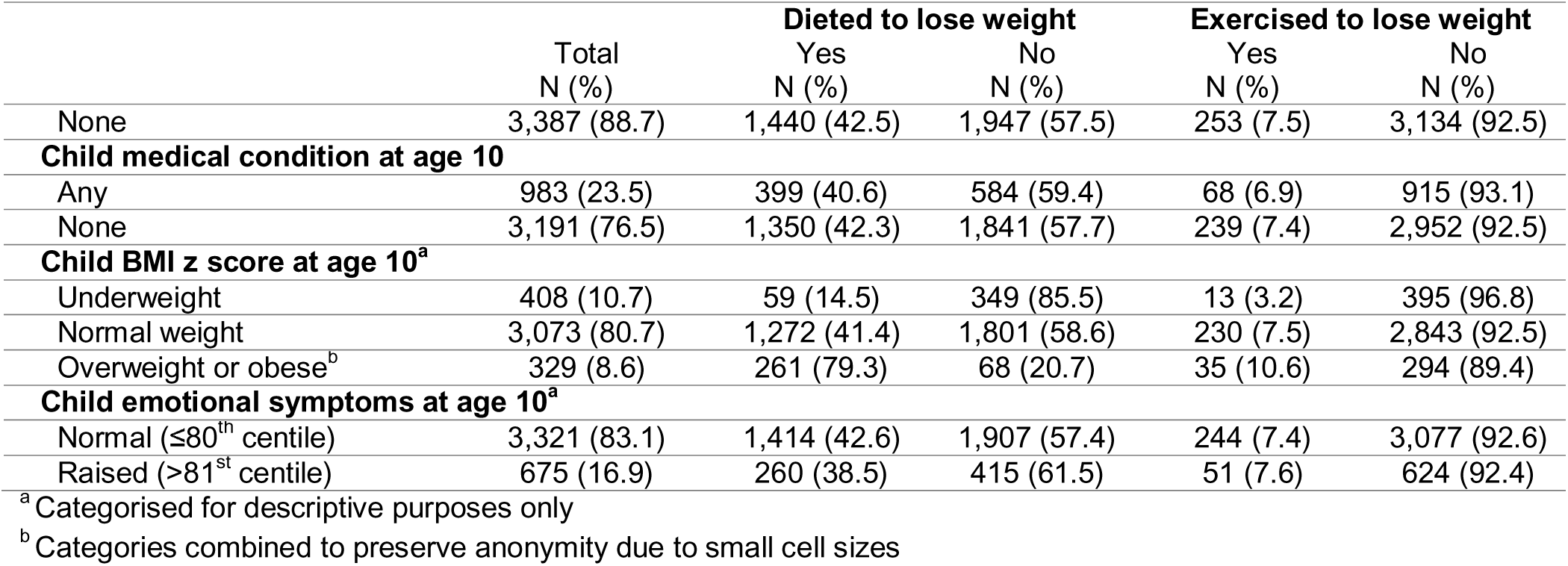
Characteristics of sample with valid exposure data (N=4,650)

|  | Total<br>N (%) | Dieted to lose weight |  | Exercised to lose weight |  |
| --- | --- | --- | --- | --- | --- |
|  |  | Yes<br>N (%) | No<br>N (%) | Yes<br>N (%) | No<br>N (%) |
| Total % | 4,650 (100) | 1,938 (41.7) | 2,712 (58.3) | 343 (7.4) | 4,307 (92.6) |
| **Child sex** |  |  |  |  |  |
| Male | 2,013 (43.3) | 354 (17.6) | 1,659 (82.4) | 102 (5.1) | 1,911 (94.9) |
| Female | 2,637 (56.7) | 1,584 (60.1) | 1,053 (39.9) | 241 (9.1) | 2,396 (90.9) |
| **Child ethnicity** |  |  |  |  |  |
| White | 4,108 (97.7) | 1,719 (41.8) | 2,389 (58.2) | 293 (7.1) | 3,815 (92.9) |
| Ethnic minority | 95 (2.3) | 41 (43.2) | 54 (56.8) | 15 (15.8) | 80 (84.2) |
| **Parent's education** |  |  |  |  |  |
| Compulsory | 3,091 (66.9) | 1,289 (41.7) | 1,802 (58.3) | 235 (7.6) | 2,856 (92.4) |
| Beyond compulsory | 1,529 (33.1) | 634 (41.5) | 895 (58.5) | 104 (6.8) | 1,425 (93.2) |
| **Father's occupational class** |  |  |  |  |  |
| Non-Manual | 1,578 (35.3) | 659 (41.8) | 919 (58.2) | 103 (6.5) | 1,475 (93.5) |
| Manual | 2,907 (64.7) | 1,194 (41.1) | 1,713 (58.9) | 227 (7.8) | 2,680 (92.2) |
| **Parents' marital status** |  |  |  |  |  |
| Married | 4,415 (95.1) | 1,823 (41.3) | 2,592 (58.7) | 326 (7.4) | 4,089 (92.6) |
| Not Married | 228 (4.9) | 111 (48.7) | 117 (51.3) | 17 (7.5) | 211 (92.5) |
| **Child birthweight<sup>a</sup>** |  |  |  |  |  |
| Low (<2500g) | 214 (4.6) | 91 (42.5) | 123 (57.5) | 17 (7.9) | 197 (92.1) |
| Normal (≥ 2500g) | 4,428 (95.4) | 1,845 (41.7) | 2,583 (58.3) | 324 (7.3) | 4,104 (92.7) |
| **Mother's age at child's birth** |  |  |  |  |  |
| <20 years | 371 (8.0) | 163 (43.9) | 208 (56.1) | 24 (6.5) | 347 (93.5) |
| 20-24 years | 1,643 (35.5) | 684 (41.6) | 959 (58.4) | 129 (7.9) | 1,514 (92.1) |
| 25-29 years | 1,523 (32.9) | 622 (40.8) | 901 (59.2) | 109 (7.2) | 1,414 (92.8) |
| 30-34 years | 730 (15.8) | 303 (41.5) | 427 (58.5) | 60 (8.2) | 670 (91.8) |
| 35+ years | 361 (7.8) | 157 (43.5) | 204 (56.5) | 20 (5.5) | 341 (94.5) |
| **Mother's smoking in pregnancy** |  |  |  |  |  |
| Non-smoker | 2,080 (44.9) | 809 (38.9) | 1,271 (61.1) | 139 (6.7) | 1,941 (93.3) |
| Stopped pre-pregnancy | 628 (13.6) | 271 (43.2) | 357 (56.8) | 43 (6.9) | 585 (93.1) |
| Smoked in pregnancy | 1,924 (41.5) | 853 (44.3) | 1,071 (55.6) | 160 (8.3) | 1,764 (91.7) |
| **Mother's drinking in pregnancy** |  |  |  |  |  |
| None | 1,983 (50.1) | 807 (40.7) | 1,176 (59.3) | 156 (7.9) | 1,827 (92.1) |
| Any | 1,972 (49.9) | 858 (43.5) | 1,114 (56.5) | 130 (6.6) | 1,842 (93.4) |
| **Mother's BMI when child aged 10<sup>a</sup>** |  |  |  |  |  |
| BMI <25 | 3,171 (77.2) | 1,240 (39.1) | 1,931 (60.9) | 220 (6.9) | 2,951 (93.1) |
| BMI ≥25 | 937 (22.8) | 477 (50.9) | 460 (49.1) | 79 (8.4) | 858 (91.6) |
| **Mother's depressive symptoms when child aged 10<sup>a</sup>** |  |  |  |  |  |
| Normal (≤80<sup>th</sup> centile) | 3,281 (83.7) | 1,357 (41.4) | 1,924 (58.6) | 237 (7.2) | 3,044 (92.8) |
| Raised (>81<sup>st</sup> centile) | 639 (16.3) | 284 (44.4) | 355 (55.6) | 53 (8.3) | 586 (91.7) |
| **Child self-esteem at age 10<sup>a</sup>** |  |  |  |  |  |
| Lowest third | 1,344 (37.4) | 623 (46.4) | 721 (53.6) | 103 (7.7) | 1,241 (92.3) |
| Middle third | 1,102 (30.6) | 478 (43.4) | 624 (56.6) | 90 (8.2) | 1,012 (91.8) |
| Highest third | 1,149 (32.0) | 398 (34.6) | 751 (65.4) | 73 (6.4) | 1,076 (93.6) |
| **Child breakfast consumption at age 10** |  |  |  |  |  |
| Everyday | 3,015 (80.4) | 1,221 (40.5) | 1,794 (59.5) | 202 (6.7) | 2,813 (93.3) |
| Sometimes or never<sup>b</sup> | 737 (19.6) | 352 (47.8) | 385 (52.2) | 74 (10.0) | 663 (90.0) |
| **Child physical activity at age 10** |  |  |  |  |  |
| Never or hardly ever | 341 (8.2) | 175 (51.3) | 166 (48.7) | 25 (7.3) | 316 (92.7) |
| Sometimes | 1,693 (40.5) | 823 (48.6) | 877 (51.4) | 148 (8.7) | 1,545 (91.3) |
| Often | 2,142 (51.3) | 749 (35.0) | 1,393 (65.0) | 136 (6.4) | 2,006 (93.6) |
| **Child TV watching at age 10** |  |  |  |  |  |
| Sometimes or never<sup>b</sup> | 898 (21.4) | 405 (45.1) | 493 (54.9) | 72 (8.0) | 826 (92.0) |
| Often | 3,293 (78.6) | 1,351 (41.0) | 1,942 (59.0) | 237 (7.2) | 3,056 (92.8) |
| **Child smoking at age 10** |  |  |  |  |  |
| Any | 431 (11.3) | 159 (36.9) | 272 (63.1) | 31 (7.2) | 400 (92.8) |
|  |  | Yes<br>N (%) | No<br>N (%) | Yes<br>N (%) | No<br>N (%) |
| None | 3,387 (88.7) | 1,440 (42.5) | 1,947 (57.5) | 253 (7.5) | 3,134 (92.5) |
| **Child medical condition at age 10** |  |  |  |  |  |
| Any | 983 (23.5) | 399 (40.6) | 584 (59.4) | 68 (6.9) | 915 (93.1) |
| None | 3,191 (76.5) | 1,350 (42.3) | 1,841 (57.7) | 239 (7.4) | 2,952 (92.5) |
| **Child BMI z score at age 10<sup>a</sup>** |  |  |  |  |  |
| Underweight | 408 (10.7) | 59 (14.5) | 349 (85.5) | 13 (3.2) | 395 (96.8) |
| Normal weight | 3,073 (80.7) | 1,272 (41.4) | 1,801 (58.6) | 230 (7.5) | 2,843 (92.5) |
| Overweight or obese<sup>b</sup> | 329 (8.6) | 261 (79.3) | 68 (20.7) | 35 (10.6) | 294 (89.4) |
| **Child emotional symptoms at age 10<sup>a</sup>** |  |  |  |  |  |
| Normal ($\leq 80^{\text{th}}$ centile) | 3,321 (83.1) | 1,414 (42.6) | 1,907 (57.4) | 244 (7.4) | 3,077 (92.6) |
| Raised ($> 81^{\text{st}}$ centile) | 675 (16.9) | 260 (38.5) | 415 (61.5) | 51 (7.6) | 624 (92.4) |

<sup>a</sup> Categorised for descriptive purposes only
<sup>b</sup> Categories combined to preserve anonymity due to small cell sizes

### Missing data

Compared to those with available data, a greater proportion of those with missing data on weight control behaviours, were boys, from disadvantaged backgrounds, smoked or had medical conditions at age 10, and had mothers who smoked in pregnancy (**eTable2**). Within the analytic sample, 4,302 (92.5%) and 4,258 (91.6%) had at least one valid measure of depressive symptom and BMI outcome data, respectively, of which about half had complete or near complete outcome data (i.e., 53.1% and 49.8% missing no or only one measure of depressive symptoms and BMI respectively between 26 and 51 years). A greater proportion of boys and children from minoritised ethnic groups, whose parents were not married, and who had mothers who smoked in pregnancy were missing outcome data at all time points (**eTable3**).

### Trajectories of depressive symptoms and BMI

There was evidence that depressive symptoms increased linearly with age, but no evidence of non-linear changes over time. BMI also increased with age, with additional evidence that the increase reduced over time. Therefore, we retained a non-linear age indicator for BMI, but not for depressive symptoms analyses. Full results in **eTable4**.

#### Association between dieting and exercise to lose weight and depression

In unadjusted analyses of our imputed sample, there was strong evidence that adolescents who had dieted (intercept mean difference (MD) 0.34, 95% CI 0.25 to 0.43) and those who exercised (MD 0.26, 95% CI 0.09 to 0.43) had higher adult depressive symptom trajectories (**Table 3**). These associations were attenuated after adjustment for child sex and ethnicity (dieting adjusted mean difference (aMD) 0.13, 95% CI 0.04 to 0.23; exercise aMD 0.18, 95% CI 0.02 to 0.35), with only weak evidence of association persisting after adjustment for family socioeconomic position, perinatal factors, mother’s BMI and depressive symptoms and child’s self-esteem, health-related behaviours and general health at age 10 (dieting aMD 0.08, 95% CI -0.01 to 0.18; exercise aMD 0.16, 95% CI 0.00 to 0.33). After the inclusion of child BMI and emotional symptoms at age 10, there was evidence that adolescents who dieted and exercised had, on average, 0.13 (95% CI 0.03 to 0.23) and 0.17 (95% CI 0.01 to 0.34) higher depressive symptom scores, respectively. There was no evidence that either association differed by sex (dieting-sex interaction p value 0.574; exercise-sex interaction p value 0.456) or linear age (dieting-age interaction p value 0.846; exercise-age interaction p value 0.656). Trajectories from final models are shown in **Figure 1A**.

**Figure 1.**
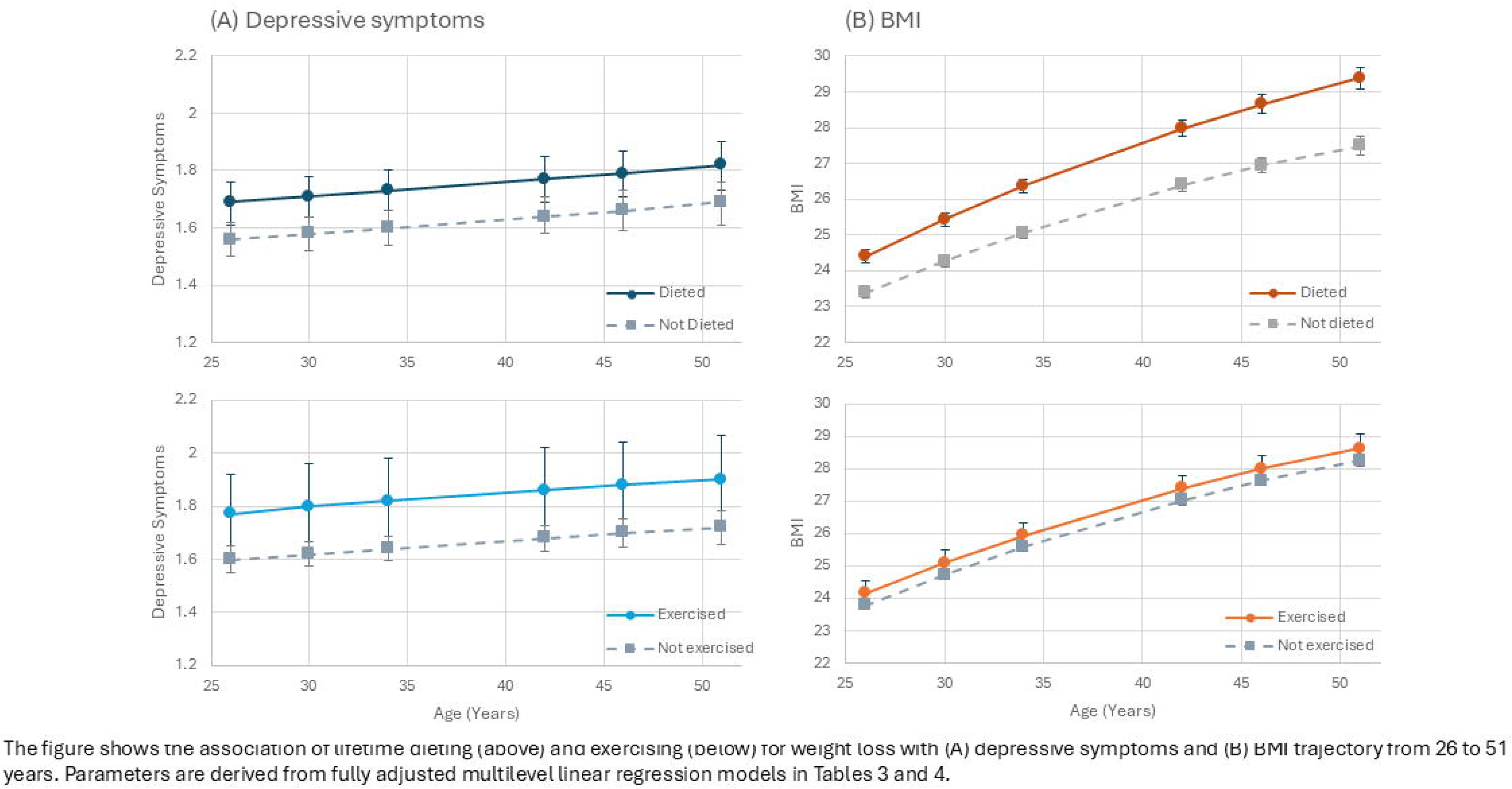
Association of weight control behaviours at 16 years with depressive symptoms and BMI from 26to 51 years.

**Table 3.** Association between dieting and exercise to lose weight at age 16 and trajectories of depressive symptoms from age 26 to 51. Results from multilevel modelling of a sample with valid exposure data and imputed outcome and confounder data (N=4,650).

| Weight control at 16 years | Mean difference in depressive symptom trajectory<sup>a</sup><br>(95% CI)<br>P value |  |  |  |  |  |  |  | Final model interaction tests<br>P value<br>Exposure x |  |
| --- | --- | --- | --- | --- | --- | --- | --- | --- | --- | --- |
|  | Model 0<sup>b</sup> | Model 1 | Model 2 | Model 3 | Model 4 | Model 5 | Model 6 | Model 7 | sex | age |
| **Dieting to lose weight** |  |  |  |  |  |  |  |  |  |  |
| Dieted (yes vs no) | 0.34<br>(0.25, 0.43)<br><0.001 | 0.13<br>(0.04, 0.23)<br>0.009 | 0.13<br>(0.03, 0.23)<br>0.009 | 0.13<br>(0.03, 0.22)<br>0.010 | 0.11<br>(0.02, 0.21)<br>0.023 | 0.08<br>(-0.01, 0.18)<br>0.093 | 0.13<br>(0.02, 0.23)<br>0.018 | 0.13<br>(0.03, 0.23)<br>0.015 | 0.574 | 0.846 |
| **Exercise to lose weight** |  |  |  |  |  |  |  |  |  |  |
| Exercised (yes vs no) | 0.26<br>(0.09, 0.43)<br>0.002 | 0.18<br>(0.02, 0.35)<br>0.032 | 0.17<br>(0.01, 0.34)<br>0.041 | 0.17<br>(0.01, 0.34)<br>0.042 | 0.17<br>(0.00, 0.33)<br>0.044 | 0.16<br>(0.00, 0.33)<br>0.049 | 0.17<br>(0.01, 0.34)<br>0.036 | 0.17<br>(0.01, 0.34)<br>0.036 | 0.456 | 0.656 |

<sup>a</sup>This is the mean difference in the intercept
<sup>b</sup>Model 0: age
Model 1: 0 + child sex and ethnicity
Model 2: 1 + parent education, father's occupational class, mother's marital status
Model 3: 2 + child birthweight, mother's age at child's birth, mother's smoking and drinking in pregnancy
Model 4: 3 + mother's BMI and depressive symptoms when child aged 10
Model 5: 4 + child self-esteem, breakfast consumption, physical activity, television watching, smoking and general health at age 10
Model 6: 5 + child BMI z score at age 10
Model 7: 6 + child emotional symptoms at age 10

#### Association between dieting and exercise to lose weight and BMI

In unadjusted analyses of our imputed sample, there was strong evidence that adolescents who had dieted (MD 1.70, 95% CI 1.45 to 1.94) and those who exercised (MD 0.92, 95% CI 0.44 to 1.39) had higher BMI trajectories (**Table 4**). The magnitude of both associations increased after adjusting for child sex and ethnicity (dieting aMD 2.70, 95% CI 2.44 to 2.96; exercise aMD 1.12, 95% CI 0.65 to 1.59). These associations were attenuated after adjusting for family socioeconomic position, perinatal factors and mother’s BMI and depressive symptoms (dieting aMD 2.42, 95% CI 2.16 to 2.67; exercise aMD 0.97, 95% CI 0.53 to 1.42). There was little further attenuation after adjusting for child self-esteem, health-related behaviours and general health at age 10. After including child BMI and emotional symptoms at age 10, BMI trajectories for those who dieted and exercised were 1.10kg/m^2^ (95% 0.83 to 1.36) and 0.37kg/m^2^ (95% CI -0.04 to 0.78) higher respectively, compared to those who had not, though evidence for the latter was weak. There was no evidence that these associations differed by sex (dieting-sex interaction p value 0.234; exercise-sex interaction p value 0.130). There was strong evidence that dieting only was associated with an increased rate of change in BMI over time (dieting-linear age p value <0.001). There was no evidence for an interaction with quadratic age for dieting when we included both dieting-linear age and dieting-quadratic age interactions together (dieting-linear age p value 0.314. dieting-quadratic age p value 0.201). Adolescents who dieted gained an additional 0.03kg/m^2^ per year (95% CI 0.02, 0.05) than those who had not dieted for weight loss, and no evidence of differences by age for exercise (exercise-linear age p value 0.958, exercise-quadratic age p value 0.915). BMI trajectories from final models are shown in **Figure 1B**.

**Table 4.** Association between dieting and exercising for weight loss at age 16 and trajectories of BMI from age 26 to 51. Results from multilevel modelling of a sample with valid exposure data and imputed outcome and confounder data (N=4,650).

| Weight control at 16 years | Mean difference in BMI trajectory<sup>a</sup><br>(95% CI)<br>P value |  |  |  |  |  |  |  | Final model interaction tests<br>P value<br>Exposure x |  |  |
| --- | --- | --- | --- | --- | --- | --- | --- | --- | --- | --- | --- |
|  | Model 0<sup>b</sup> | Model 1 | Model 2 | Model 3 | Model 4 | Model 5 | Model 6 | Model 7 | sex | age | age<sup>2</sup> |
| **Dieting to lose weight** |  |  |  |  |  |  |  |  |  |  |  |
| Dieted (yes vs no) | 1.70<br>(1.45, 1.94)<br><0.001 | 2.70<br>(2.44, 2.96)<br><0.001 | 2.71<br>(2.45, 2.97)<br><0.001 | 2.68<br>(2.42, 2.93)<br><0.001 | 2.42<br>(2.16, 2.67)<br><0.001 | 2.41<br>(2.15, 2.66)<br><0.001 | 1.09<br>(0.83, 1.35)<br><0.001 | 1.10<br>(0.83, 1.36)<br><0.001 | 0.234 | <0.001 | 0.314<br>0.201 |
| **Exercise to lose weight** |  |  |  |  |  |  |  |  |  |  |  |
| Exercised (yes vs no) | 0.92<br>(0.44, 1.39)<br><0.001 | 1.12<br>(0.65, 1.59)<br><0.001 | 1.09<br>(0.63, 1.56)<br><0.001 | 1.04<br>(0.57, 1.50)<br><0.001 | 0.97<br>(0.53, 1.42)<br><0.001 | 0.96<br>(0.52, 1.42)<br><0.001 | 0.37<br>(-0.04, 0.78)<br>0.079 | 0.37<br>(-0.04, 0.78)<br>0.079 | 0.130 | 0.923 | 0.958<br>0.915 |

<sup>a</sup> This is the mean difference in the intercept
<sup>b</sup> Model 0: age, age<sup>2</sup>
Model 1: 0 + child sex and ethnicity
Model 2: 1 + parent education, father's occupational class, mother's marital status
Model 3: 2 + child birthweight, mother's age at child's birth, mother's smoking and drinking in pregnancy
Model 4: 3 + mother's BMI and depressive symptoms when child aged 10
Model 5: 4 + child self-esteem, breakfast consumption, physical activity, television watching, smoking and general health at age 10
Model 6: 5 + child BMI z score at age 10
Model 7: 6 + child emotional symptoms at age 10

#### Sensitivity analyses

Results of analyses among participants with complete data were consistent with those of the main analyses (**eTables 5-6**). Similarly, results of analyses using self-reported vs objectively measured BMI at age 46 were comparable (**eTable7**).

## Discussion

Depressive symptoms and BMI increased over the lifecourse. Compared to those who did not diet, among those who dieted, trajectories of depressive symptoms were higher at all time points and those of BMI were higher and increasing. Those who exercised also had higher trajectories of depressive symptoms at all time points.

These associations remained after adjustment for prior BMI and emotional difficulties. We found weak evidence of differences in adult BMI trajectory among those who had exercised to lose weight at age 16.

We note several limitations. First, measures of dieting and exercise came from self-reported lifetime behaviours, which could have limited our ability to establish temporality when controlling for prior differences in BMI and emotional difficulties at age 10, although these are likely to precede attempts to lose weight for most participants.^23^ Attempts to lose weight were captured as binary indicators, without further information on how these were carried out, such as whether following a structured, doctor- or dietician-supervised diet or using unhealthy weight control behaviours, which may have different associations with depressive symptoms and BMI.^14,25^

Second, measures of BMI came from self-reported height and weight, which may be more prone to reporting bias. However, our sensitivity analyses suggested consistency between associations for self-reported and nurse-measured BMI at age 46.

Finally, although we controlled for a wide range of confounders, in observational studies there may remain residual confounding due to imperfectly measured, unmeasured or unknown factors.

Our results are in line with previous longitudinal evidence^7–13^, extending this to suggest that associations between weight control behaviours and higher depressive symptoms and BMI may persist into mid-life. Although the magnitude of associations with depressive symptoms in our study was relatively small at an individual level, a small shift in the distribution of depressive symptom scores at the population level is likely to result in large numbers of new depression cases, particularly given the high prevalence of dieting. In contrast to some studies^6,7^, we did not find evidence that this association differed in girls and boys. Potentially, sex differences may be seen at younger ages when depression is less common among boys.

Notably, we found adolescents who dieted gained increasingly more weight over adulthood than non-dieters. This could reflect long-term physiological adaptations to weight cycling or “yo-yo dieting”^26^ (an individual initially loses weight, then regains all or more weight after stopping their diet). Previous research suggests weight control behaviours may progress into disordered eating^10^, and both weight control and disordered eating behaviours in adolescence are likely to track into adulthood.^27^ Thus, dieting in adolescence may develop into long-term weight cycling, which increases the risk of cardiovascular disease^28^ and diabetes.^29^

Though our findings reflect lifecourse impacts of dieting in the 1980s, they raise important questions about the adult consequences of pressures to lose weight among more recent adolescents. Current UK public health strategies targeting childhood obesity have emphasised individual responsibility; school lessons teach about the importance of calorie restriction and exercise to prevent obesity, while calories on menus and nutritional labelling aim to encourage healthy choices, with little evidence of effectiveness.^30–32^ Evaluation of the potential unintended consequences of these strategies is necessary as, if causal, our findings suggest that reducing pressures to lose weight in adolescence may help promote better lifecourse mental and physical health.

## Data Availability

Data are available from the UK Data Service

https://datacatalogue.ukdataservice.ac.uk/series/series/200001#abstract

