## Supplement for "Adolescent weight control behaviours and adult depressive symptom and body mass index trajectories"

**Supplementary material**

**eTable 1.** Items in the Malaise Inventory in the full scale and the nine-item scale (in bold).

| **Item from full scale** | **Included in nine-item scale?** |
| --- | --- |
| 1. Do you often have backache? |  |
| 2. **Do you feel tired most of the time?** | **Yes** |
| 3. **Do you often feel depressed?** | **Yes** |
| 4. Do you often have bad headaches? |  |
| 5. **Do you often get worried about things?** | **Yes** |
| 6. Do you usually have great difficulty in falling or staying asleep? |  |
| 7. Do you usually wake unnecessarily early in the morning? |  |
| 8. Do you wear yourself out worrying about your health? |  |
| 9. **Do you often get into a violent rage?** | **Yes** |
| 10. Do people annoy and irritate you? |  |
| 11. Have you at times had a twitching of the face, head or shoulders? |  |
| 12. **Do you suddenly become scared for no good reason?** | **Yes** |
| 13. Are you scared to be alone when there are not friends near you? |  |
| 14. **Are you easily upset or irritated?** | **Yes** |
| 15. Are you frightened of going out alone or of meeting people? |  |
| 16. **Are you constantly keyed up and jittery?** | **Yes** |
| 17. Do you suffer from indigestion? |  |
| 18. Do you suffer from an upset stomach? |  |
| 19. Is your appetite poor? |  |
| 20. **Does every little thing get on your nerves and wear you out?** | **Yes** |
| 21. **Does your heart often race like mad?** | **Yes** |
| 22. Do you often have bad pain in eyes? |  |
| 23. Are you troubled with rheumatism or fibrosis? |  |
| 24. Have you ever had a nervous breakdown? |  |

**eMethod 1.** Confounding variables

We adjusted for the following theoretically informed confounders:

- Child sex at birth (male versus female) – taken from maternity notes as part of the first sweep
- Child ethnicity (White versus ethnic minority) – reported as “English, Welsh, Scottish, Northern Irish”, “Irish”, “Other European”, all coded as White, or “West Indian or Guyanese”, “Indian”, “Pakistani”, “Bangladeshi”, “Mixed parentage or any other ethnic group”, which were all coded as ethnic minority. These were reported by their parent when the child was aged 10. We chose to group any White ethnic groups in this way to capture their similarity to Eurocentric beauty standards, though we acknowledge that Irish children were likely to have experienced discrimination during this time.
- Highest parent education (compulsory versus beyond compulsory education) – parents reported the age at which they left education. For each parent, we derived a binary variable indicating whether they had stayed in education beyond age 15, which was the school leaving age from 1947 to 1972. We then created a composite variable taking the higher value of parents in the household in order to incorporate data from both parents in two-parent households. In single-parent households, this composite variable took the value of the present parent.
- Father’s occupational class (manual versus non-manual) – in the first sweep, data were collected on the occupational class of the child’s father and coded according to the Register General’s classification. Classifications coded as manual were Class IIIM, Class IV and Class V. Classifications coded as non-manual were Class I, Class II and Class IIIN.
- Marital status (married versus unmarried) – this was reported by the child’s mother in the first sweep as: single, married, separated, divorced, widowed. Any marital status except married was coded as unmarried.
- Child birthweight – this was taken from the first sweep. For the purposes of descriptive analyses only, we derived a new variable indicating whether the child was born with a low birthweight (<2500g) or not (≥2500g).
- Mother’s age at child’s birth – this was reported by mothers in the first sweep and coded into 4-year age bands: under 20 years; 20-24 years; 25-29 years; 30-34 years; 35 years and older. We used a categorical age variable as we expected that mothers’ age would be non-linearly associated with child health.
- Mother’s cigarette use in pregnancy – in the first sweep, mothers were asked whether they currently smoked, if they had ever smoked, and if they had smoked during their pregnancy. From these data, we derived a new variable to indicate whether they never smoked; stopped before pregnancy; or smoked during pregnancy.
- Mother’s alcohol use in pregnancy (none versus any) – these data were retrospectively reported by mothers when their child was aged 10 as frequency of consuming an alcoholic drink, both during early and late pregnancy (Most days; 2-3 times a week; Once a week or less; Not at all). We used the early pregnancy measure.
- Mother’s BMI – this was calculated from self-reported height and weight when their child was aged 10. For descriptive purposes only, we created a categorical variable to indicate whether their BMI was < 25kg/m^2^ or ≥25kg/m^2^.
- Mother’s depressive symptoms – when their child was aged 10, mothers completed the Malaise Inventory, reporting their own experiences of depressive symptoms. Responses to all items were summed, with higher scores indicate greater depressive symptoms. For descriptive purposes only, we used a categorical variable derived by the BCS70 team which indicated whether their Malaise score was normal (those ≤80^th^ centile) or raised (81^st^ centile or above) based on the distribution.
- Child self-esteem – this was self-reported by the child at age 10 using the Lawrence Self-Esteem Questionnaire (LAWSEQ), a 16-item questionnaire capturing perceived self-competency, social exclusion and shyness. We created a sum score of responses to all items where higher scores indicate greater self-esteem. For descriptive purposes only, we created a categorical variable based on tertiles of self-esteem scores.
- Child breakfast consumption – child participants were asked when they were aged 10 whether they have something to eat before coming to school in the morning, with possible options “Yes always”, “Sometimes”, “No never”.
- Child physical activity – mothers were asked when their child was aged 10 how often their child plays sports in their spare time (Never or hardly ever, Sometimes, Often).
- Child television watching – mothers were asked when their child was aged 10 how often their child watches television in their spare time (Never or hardly ever, Sometimes, Often). We included this as it may index sedentary behaviour and exposure to beauty standards in televisual media.
- Child smoking – at age 10, child participants were asked if they had ever tried a cigarette (Yes or No).
- Child’s general health – when their child was aged 10, their mother was asked whether the child has any medical condition or illness, behavioural problem or educational difficulty which they consider to be important (Yes or No).
- Child’s BMI z score at age 10 – medical staff measured the height and weight of the child when they were aged 10. We derived age- and sex-standardised scores of their BMI according to British 1990 reference values. For descriptive purposes only, we categorised their BMI z scores according to International Obesity Task Force cut offs.
- Child’s emotional difficulties at age 10 – these were reported by their mother using the Rutter Behaviour Scale, which captures emotional symptoms and behavioural difficulties, as well as peer problems, on a visual-analogue scale. Responses to all items were summed, with higher scores indicating greater difficulties. For descriptive purposes only, we used a categorical variable derived by the BCS70 team based on the distribution of scores (normal i.e., scores ≤80^th^ centile; versus raised, i.e., 81^st^ centile and above).

**eMethod2.** Multiple Imputation

Auxiliary variables in multiple imputation were measures of child BMI and emotional difficulties at age 5, father’s BMI when the child was aged 10, and nurse-measured BMI at age 46 for the child cohort member. We conducted multiple imputation by chained equations in Stata version 18.5 using the command *mi impute chained*, generating 50 datasets. The imputation model included all exposures, outcomes and confounders, alongside auxiliary variables. These were measures of child BMI and emotional difficulties at age 5, father’s BMI when the child was aged 10, and nurse-measured BMI at age 46 for the child cohort member. To appropriately account for nested structure of the data (repeated outcome measures within individuals), we reshaped the data in wide format before imputation and subsequently reshaped into long format for multilevel modelling.

**eTable 2.** Comparison of full cohort and analytic sample characteristics and frequencies and proportions of missing data among those with valid exposure data. Proportions of valid data add up to 100%; missing data proportions calculated separately as a proportion of total analytic sample.

|  | Full cohort | Sample with valid exposure |
| --- | --- | --- |
| Total N (%) | 16,672 (100) | 4,650 (100) |
| Child sex |  |  |
| Male | 8,632 (51.8) | 2,013 (43.3) |
| Female | 8,040 (48.2) | 2,637 (56.7) |
| *Missing* |  | 0 |
| Child ethnicity |  |  |
| White | 12,153 (96.9) | 4,108 (97.7) |
| Ethnic minority | 394 (3.1) | 95 (2.3) |
| *Missing* |  | 447 (9.6) |
| Parent’s education |  |  |
| Compulsory | 12,099 (73.1) | 3,091 (66.9) |
| Beyond compulsory | 4,457 (26.9) | 1,529 (33.1) |
| *Missing* |  | 30 (0.7) |
| Father’s occupational class |  |  |
| Non-Manual | 4,516 (28.6) | 1,578 (35.3) |
| Manual | 11,273 (71.4) | 2,907 (64.7) |
| *Missing* |  | 165 (3.5) |
| Parents’ marital status |  |  |
| Married | 15,440 (92.7) | 4,415 (95.1) |
| Not Married | 1,219 (7.3) | 228 (4.9) |
| *Missing* |  | 7 (0.2) |
| Child birthweight |  |  |
| Low (<2500g) | 1,073 (6.4) | 214 (4.6) |
| Normal (≥ 2500g) | 15,583 (93.6) | 4,428 (95.4) |
| *Missing* |  | 8 (0.2) |
| Mother’s age at child’s birth |  |  |
| <20 years | 1,628 (9.8) | 371 (8.0) |
| 20-24 years | 5,929 (35.8) | 1,643 (35.5) |
| 25-29 years | 5,099 (30.8) | 1,523 (32.9) |
| 30-34 years | 2,520 (15.2) | 730 (15.8) |
| 35+ years | 1,398 (8.4) | 361 (7.8) |
| *Missing* |  | 22 (0.5) |
| Mother’s smoking in pregnancy |  |  |
| Non-smoker | 6,991 (42.1) | 2,080 (44.9) |
| Stopped pre-pregnancy | 1,975 (11.9) | 628 (13.6) |
| Smoked in pregnancy | 7,629 (46.0) | 1,924 (41.5) |
| *Missing* |  | 18 (0.4) |
| Mother’s drinking in pregnancy |  |  |
| None | 6,112 (52.2) | 1,983 (50.1) |
| Any | 5,614 (47.8) | 1,972 (49.9) |
| *Missing* |  | 695 (14.9) |
| Mother’s BMI when child aged 10 |  |  |
| BMI <25 | 9,204 (75.4) | 3,171 (77.2) |
| BMI ≥25 | 3,006 (24.6) | 937 (22.8) |
| *Missing* |  | 542 (11.7) |
| Mother’s depressive symptoms when child aged 10 |  |  |
| Normal (≤80^th^ centile) | 9,211 (80.2) | 3,281 (83.7) |
| Raised (>81^st^ centile) | 2,277 (19.8) | 639 (16.3) |
| *Missing* |  | 730 (15.7) |

|  | Full cohort | Sample with valid exposure |
| --- | --- | --- |
| Child self-esteem at age 10 |  |  |
| Lowest third | 4,247 (39.4) | 1,344 (37.4) |
| Middle third | 3,320 (30.8) | 1,102 (30.6) |
| Highest third | 3,207 (29.8) | 1,149 (32.0) |
| *Missing* |  | 1,055 (22.7) |
| Child breakfast consumption at age 10 |  |  |
| Everyday | 8,823 (78.6) | 3,015 (80.4) |
| Sometimes or never | 2,408 (21.4) | 737 (19.6) |
| *Missing* |  | 898 (19.3) |
| Child physical activity at age 10 |  |  |
| Never or hardly ever | 969 (7.8) | 341 (8.2) |
| Sometimes | 4,661 (37.6) | 1,693 (40.5) |
| Often | 6,783 (54.6) | 2,142 (51.3) |
| *Missing* |  | 474 (10.2) |
| Child TV watching at age 10 |  |  |
| Sometimes or never | 2,576 (20.7) | 898 (21.4) |
| Often | 9,852 (79.3) | 3,293 (78.6) |
| *Missing* |  | 459 (9.9) |
| Child smoking at age 10 |  |  |
| Any | 1,611 (14.0) | 431 (11.3) |
| None | 9,862 (86.0) | 3,387 (88.7) |
| *Missing* |  | 832 (17.9) |
| Child medical condition at age 10 |  |  |
| Any | 3,310 (26.7) | 983 (23.5) |
| None | 9,085 (73.3) | 3,191 (76.5) |
| *Missing* |  | 476 (10.2) |
| Child BMI z score at age 10 |  |  |
| Underweight | 1,196 (10.8) | 408 (10.7) |
| Normal weight | 8,921 (80.2) | 3,073 (80.7) |
| Overweight or obese | 999 (9.0) | 329 (8.6) |
| *Missing* |  | 840 (18.1) |
| Child emotional symptoms at age 10 |  |  |
| Normal (≤80^th^ centile) | 9,355 (80.0) | 3,321 (83.1) |
| Raised (>81^st^ centile) | 2,343 (20.0) | 675 (16.9) |
| *Missing* |  | 654 (14.1) |

**eTable 3.** Characteristics of sample with missing outcome data, among those with valid exposure data.

|  | Missing depressive symptom data^a^ | | Missing BMI data^a^ | |
| --- | --- | --- | --- | --- |
|  | No % | Yes % | No % | Yes % |
| Total N (%) | 4302 (92.5) | 348 (7.5) | 4258 (91.6) | 392 (8.4) |
| Child sex |  |  |  |  |
| Male | 90.0 | 10.0 | 88.4 | 11.6 |
| Female | 94.5 | 5.5 | 94.0 | 6.0 |
| Child ethnicity |  |  |  |  |
| White | 93.0 | 7.0 | 92.1 | 7.9 |
| Ethnic minority | 88.4 | 11.6 | 87.4 | 12.6 |
| Parent’s education |  |  |  |  |
| Compulsory | 91.7 | 8.3 | 90.7 | 9.3 |
| Beyond compulsory | 94.2 | 5.8 | 93.3 | 6.7 |
| Father’s occupational class |  |  |  |  |
| Non-Manual | 93.9 | 6.1 | 92.9 | 7.1 |
| Manual | 91.9 | 8.1 | 91.0 | 9.0 |
| Parents’ marital status |  |  |  |  |
| Married | 89.5 | 10.5 | 87.7 | 12.3 |
| Not Married | 92.7 | 7.3 | 91.8 | 8.2 |
| Child birthweight |  |  |  |  |
| Low (<2500g) | 92.1 | 7.9 | 92.5 | 7.5 |
| Normal (≥ 2500g) | 92.6 | 7.4 | 91.5 | 8.5 |
| Mother’s age at child’s birth |  |  |  |  |
| <20 years | 92.2 | 7.8 | 90.6 | 9.4 |
| 20-24 years | 92.0 | 8.0 | 91.2 | 8.8 |
| 25-29 years | 92.6 | 7.4 | 91.8 | 8.2 |
| 30-34 years | 94.0 | 6.0 | 92.9 | 7.1 |
| 35+ years | 92.2 | 7.8 | 90.9 | 9.1 |
| Mother’s smoking in pregnancy |  |  |  |  |
| Non-smoker | 94.2 | 5.8 | 93.8 | 6.2 |
| Stopped pre-pregnancy | 93.6 | 6.4 | 91.1 | 8.9 |
| Smoked in pregnancy | 90.4 | 9.6 | 89.4 | 10.6 |
| Mother’s drinking in pregnancy |  |  |  |  |
| No | 91.6 | 8.4 | 90.7 | 9.3 |
| Yes | 94.3 | 5.7 | 93.4 | 6.6 |
| Mother’s BMI when child aged 10 |  |  |  |  |
| BMI <25 | 93.0 | 7.0 | 92.1 | 7.9 |
| BMI ≥25 | 92.3 | 7.7 | 91.6 | 8.4 |
| Mother’s depressive symptoms when child aged 10 |  |  |  |  |
| Normal (≤80^th^ centile) | 93.2 | 6.8 | 92.2 | 7.8 |
| Raised (>81^st^ centile) | 91.5 | 8.5 | 91.4 | 8.6 |
| Child self-esteem at age 10 |  |  |  |  |
| Lowest third | 92.7 | 7.3 | 91.9 | 8.1 |
| Middle third | 93.2 | 6.8 | 92.5 | 7.5 |
| Highest third | 93.6 | 6.4 | 92.2 | 7.8 |
| Child breakfast consumption at age 10 |  |  |  |  |
| Everyday | 93.4 | 6.6 | 92.3 | 7.7 |
| Sometimes or never | 92.5 | 7.5 | 92.0 | 8.0 |
| Child physical activity at age 10 |  |  |  |  |
| Never or hardly ever | 93.3 | 6.7 | 92.4 | 7.6 |
| Sometimes | 93.4 | 6.6 | 92.6 | 7.4 |
| Often | 92.4 | 7.6 | 91.4 | 8.6 |
| Child TV watching at age 10 |  |  |  |  |
| Sometimes or never | 94.3 | 5.7 | 93.5 | 6.5 |
| Often | 92.5 | 7.5 | 91.5 | 8.5 |
| Child smoking at age 10 |  |  |  |  |
| Yes | 93.2 | 6.8 | 92.4 | 7.6 |
| No | 92.6 | 7.4 | 90.7 | 8.3 |
| Child medical condition at age 10 |  |  |  |  |
| Yes | 93.2 | 6.8 | 92.2 | 7.8 |
| No | 91.8 | 8.2 | 91.2 | 8.8 |
| Child BMI z score at age 10 |  |  |  |  |
| Underweight | 92.4 | 7.6 | 90.2 | 9.8 |
| Normal weight | 93.0 | 7.0 | 92.1 | 7.9 |
| Overweight or obese | 94.2 | 5.8 | 93.3 | 6.7 |
| Child emotional symptoms at age 10 |  |  |  |  |
| Normal (≤80^th^ centile) | 93.4 | 6.6 | 92.5 | 7.5 |
| Raised (>81^st^ centile) | 90.7 | 9.3 | 90.2 | 9.8 |

^a^ Participants with at least one valid measure of each outcome coded as not missing (no column); those with missing outcome data at all follow-up sweeps coded as missing (yes column).

**eTable 4.** Mean depressive symptom and BMI scores from ages 26 to 51

|  | Age 26 | Age 30 | Age 34 | Age 42 | Age 46 | Age 51 |
| --- | --- | --- | --- | --- | --- | --- |
| Mean depressive  symptoms (95% CI) | 1.61  (1.57, 1.66) | 1.63  (1.59, 1.68) | 1.65  (1.61, 1.70) | 1.70  (1.65, 1.74) | 1.72  (1.66, 1.77) | 1.74  (1.68, 1.80) |
| Mean BMI  (95% CI) | 23.8  (23.7, 23.9) | 24.7  (24.6, 24.9) | 25.6  (25.5, 25.7) | 27.1  (26.9, 27.2) | 27.7  (27.5, 27.8) | 28.3  (28.1, 28.5) |

**eTable 5**. Association between dieting and exercise to lose weight at age 16 and trajectories of depressive symptoms from age 26 to 51. Results from multilevel modelling of a sample with complete exposure, outcome and confounder data (N=2,291).

| Weight  control  at 16 years | Mean difference in depressive symptom trajectory  (95% CI)  P value | | | | | | | | Final model interaction tests  P value  Exposure x | |
| --- | --- | --- | --- | --- | --- | --- | --- | --- | --- | --- |
|  | Model 0^a^ | Model 1 | Model 2 | Model 3 | Model 4 | Model 5 | Model 6 | Model 7 | sex | age |
| Dieting to lose weight | | | | | | | | |  |  |
| Dieted  (yes vs no) | 0.34  (0.22, 0.46)  <0.001 | 0.14  (0.01, 0.27)  0.034 | 0.14  (0.01, 0.26)  0.036 | 0.13  (0.01, 0.26)  0.039 | 0.12  (-0.00, 0.25)  0.060 | 0.09  (-0.04, 0.21)  0.191 | 0.16  (0.02, 0.30)  0.022 | 0.16  (0.02, 0.30)  0.021 | 0.998 | 0.948 |
| Exercise to lose weight | | | | | | | | |  |  |
| Exercised  (yes vs no) | 0.32  (0.09, 0.55)  0.006 | 0.24  (0.02, 0.47)  0.033 | 0.24  (0.01, 0.46)  0.038 | 0.23  (0.01, 0.45)  0.043 | 0.24  (0.02, 0.46)  0.033 | 0.22  (-0.00, 0.44)  0.053 | 0.24  (0.02, 0.46)  0.034 | 0.24  (0.02, 0.46)  0.035 | 0.704 | 0.736 |

^a^ Model 0: age
Model 1: 0 + child sex and ethnicity
Model 2: 1 + parent education, father’s occupational class, mother’s marital status
Model 3: 2 + child birthweight, mother’s age at child’s birth, mother’s smoking and drinking in pregnancy
Model 4: 3 + mother’s BMI and depressive symptoms when child aged 10
Model 5: 4 + child self-esteem, breakfast consumption, physical activity, television watching, smoking and general health at age 10
Model 6: 5 + child BMI z score at age 10
Model 7: 6 + child emotional symptoms at age 10

**eTable 6**. Association between dieting and exercising for weight loss at age 16 and trajectories of BMI from age 26 to 51. Results from multilevel modelling of a sample with complete exposure, outcome and confounder data (N=2,274).

| Weight  control  at 16 years | Mean difference in BMI trajectory  (95% CI)  P value | | | | | | | | Final model interaction tests  P value  Exposure x | | |
| --- | --- | --- | --- | --- | --- | --- | --- | --- | --- | --- | --- |
|  | Model 0^a^ | Model 1 | Model 2 | Model 3 | Model 4 | Model 5 | Model 6 | Model 7 | sex | age | age  age^2^ |
| Dieting to lose weight | | | | | | | | |  |  |  |
| Dieted  (yes vs no) | 1.46  (1.11, 1.82)  <0.001 | 2.53  (2.16, 2.89)  <0.001 | 2.52  (2.16, 2.89)  <0.001 | 2.51  (2.15, 2.88)  <0.001 | 2.24  (1.88, 2.59)  <0.001 | 2.23  (1.88, 2.59)  <0.001 | 1.10  (0.74, 1.46)  <0.001 | 1.11  (0.75, 1.46)  <0.001 | 0.734 | <0.001 | 0.518  0.270 |
| Exercise to lose weight | | | | | | | | |  |  |  |
| Exercised  (yes vs no) | 0.80  (0.12, 1.48)  0.022 | 1.06  (0.39, 1.73)  0.002 | 1.04  (0.37, 1.71)  0.002 | 0.98  (0.31, 1.65)  0.004 | 0.93  (0.29, 1.56)  0.004 | 0.89  (0.25, 1.53)  0.006 | 0.37  (-0.21, 0.95)  0.215 | 0.36  (-0.22, 0.94)  0.222 | 0.651 | 0.525 | 0.773  0.806 |

^a^ Model 0: age, age^2^
Model 1: 0 + child sex and ethnicity
Model 2: 1 + parent education, father’s occupational class, mother’s marital status
Model 3: 2 + child birthweight, mother’s age at child’s birth, mother’s smoking and drinking in pregnancy
Model 4: 3 + mother’s BMI and depressive symptoms when child aged 10
Model 5: 4 + child self-esteem, breakfast consumption, physical activity, television watching, smoking and general health at age 10
Model 6: 5 + child BMI z score at age 10
Model 7: 6 + child emotional symptoms at age 10

**eTable 7.** Univariable and multivariable regression models of the association between dieting and exercise for weight loss and self-reported and nurse-measured BMI at age 46. Sample with valid exposure data and imputed outcomes and confounders (N=4,650)

| Weight control at 16 | Mean difference in self-reported BMI at 46  (95% CI) | | Mean difference in nurse-measured BMI at 46  (95% CI) | |
| --- | --- | --- | --- | --- |
|  | **Unadjusted** | **Fully adjusted^a^** | **Unadjusted** | **Fully adjusted^a^** |
| Dieting for weight loss |  |  |  |  |
| Dieted (yes vs no) | 2.38 (2.01, 2.75) | 1.27 (0.84, 1.71) | 2.59 (2.20, 2.98) | 1.33 (0.87, 1.79) |
| Exercise for weight loss |  |  |  |  |
| Exercised (yes vs no) | 0.97 (0.20, 1.74) | 0.17 (-0.55, 0.90) | 1.19 (0.43, 1.94) | 0.34 (-0.35, 1.04) |

^a^ Adjusted for child sex and ethnicity; parents’ education and marital status; father’s job; mother’s age at child’s birth, smoking and drinking in pregnancy, and BMI and depressive symptoms when their child was aged 10; child birthweight; child self-esteem, breakfast consumption, physical activity, television watching, smoking, general health, BMI z score and emotional difficulties at age 10.
